# Patient reported outcome on endourethrotomy combined with Amplatz dilatation for treatment of incompletely obliterated posterior urethral stricture following pelvic trauma: Validation in the era of urethroplasty

**DOI:** 10.1101/2021.07.25.21261072

**Authors:** Mohamed Wishahi, Ahmed Mehena

## Abstract

**Introduction:** Direct vision internal urethrotomy (DVIU) is gaining revival among urologists, the procedure having the advantages of being minimally invasive as well as keeping continence and sexual function unaffected. The recurrence rate at 12 months was diminished by combination of DVIU with a Amplatz dilatation at the time of the procedure. Objectives are of present work is to present the outcome of the procedure.

**Materials and methods:** This is an observational study with a cross-sectional approach. Patients underwent combined endo-urethrotomy and Amplatz dilation for posterior urethral strictures between February 2015 and January2020 were purposively recruited. We performed a retrospective review of the records of 48 patients who completed 12 months follow-up. Patient-reported outcome measures for urethral stricture surgery (USS-PROMs) and sexual function were assessed at 12 months follow-up. A questionnaire was used to assess quality of life, patients in the study filled a questionnaire. Correlation between status before and after treatment were analyzed using <u>simple statistical analysis</u> to evaluate the effect of treatment on quality of life. Clinician-driven outcome were uroflowmetry, retrograde urethrogram, and reporting of complications.

**Results:** Mean length of stricture segment was 2 cm (range 1-2.5 cm). At 12 months follow up patients reported very satisfactory. Restenosis that was treated with redo was reported in 2%. Mild stenosis that was treated with Amplatz dilatation was in 3%. Patients reported very satisfactory or satisfactory for USS-PROSMs, sexual function in comparison to pre-trauma status.

**Conclusions:** Endourethrotomy combined with Amplatz dilation is an effective treatment for short segment passable posterior urethral stricture and it has a place in the era of endourethrotomy.

## INTRODUCTION

Urethral stricture disease (USD) is one of the common diseases affecting mostly men and having a negative impact on health-related quality of life (HRQOL). It is classified into anterior urethral stricture and posterior urethral stricture (PUS). Pelvic trauma is a cause of PUS which is attributed to a high incidence of morbidities, and urinary obstruction.

The standard treatment of PUS was DVIU. Recently, numerous studies have suggested that DVIU has a similar result as surgical urethroplasties. Sachse is credited for the invention of internal visual urethrotomy using sharp cold knife, DVIU became standard in urological practice having the advantages of being minimally invasive, having no difficulty to be repeated (1), (2).

Comparison between urethroplasty versus endoscopic urethrotomy in adult men with recurrent bulbar urethral stricture showed that both procedure improved voiding symptoms and resulted in similar and better symptom scores (3).

No significant difference in recurrence between DVIU and urethral dilatation (4). Results of DVIU for patients having recurrent bulbar urethral stricture with mean stricture length of 2.0 cm was better than those with stricture <3 cm (5).

In terms of outcome, both holmium laser and cold knife urethrotomy are safe and equally effective in treating short-segment bulbar urethral strictures, complication rate, patient-reported outcome, and clinical evaluation in long term follow-up (6).

Nomikos et al reported success of DVIU following dilatation of complex urethral stricture with Amplatz renal dilators (7).

Akkoc et al reported successful treatment of urethral stricture by urethral dilatation alone using Amplatz renal dilators only (8). Guided urethral dilation and internal urethrotomy were reported to be safe, short time procedures, and offer satisfactory results with improvement in Qmax and with no recurrence at 12 months (9).

Amplatz dilation was reported to be is a good option as the initial treatment for urethral stricture (10).The validated patient-reported outcome measure (PROM) is a validated measurement to assess patient reports on surgical procedures, it was applied for urethral stricture surgery (USS) and became USS-PROM (11).

Sexual dysfunction (SD) following urethroplasty is multifactorial and is more common after posterior urethroplasty, whereby transecting bulbar urethroplasty compared to the non-transecting procedure leads to greater sexual dysfunction(SD) (12). Sexual dysfunction causes dissatisfaction even after a successful urethral surgery (13).

Erickon et al applied the inventory proposed by O’Leary et al for assessment of sexual function before and after urethroplasty, sexual function questions on pre- and post-treatment erection and ejaculation are initiated the scores are a total of 8 points in the ejaculatory function domain, 12 in the erectile function domain, and 8 in the sexual dysfunction domain. This questionnaire compares outcomes with normative data (14),(15).

Clinician-derived outcome measurements are maximum flow rate (Qmax), and voiding cystourethrogram and/ or retrograde urethrogram.

## MATERIALS AND METHODS

We retrospectively reviewed the records of patients who had been treated for USD between February 2015 and January 2020. Of these patients, 48 had a delayed treatment of posterior urethral stricture following pelvic trauma. They were adult (>18 years), had been managed initially with suprapubic catheter (Table 1).

**Table 1.** Baseline characteristics

| Patients' demographic data |  |
| --- | --- |
| Total number | 48 |
| Mean age. years (range) | 38(20-77) |
| Mean stricture length. Cm(Range) | 2(1-2.5) |
|  | n(%) patients |
| Stricture location |  |
| Bulbar urethra | 30 ( 62.5%) |
| Membranous urethra | 18 (37.5) |
| Stricture length | 1.2.5 cm |
| Etiology; Pelvic trauma | All |
| Suprapubic catheter | All |
| Time from trauma to surgery. Months |  |
| 1-3 months | 25(52%) |
| 3-6 months | 23(48%) |
| Patient's symptoms: Inability to pass urine | All |
| Comorbidities: single/ multiple |  |
| Diabetes mellitus |  |
| Cardiovascular disease |  |
| Chronic obstructive pulmonary disease |  |
| Single comorbidity | 11(23%) |
| Multiple comorbidities; two or more | 7 (14.5%) |

The study was reviewed and approved by the ethics committee of Teodor Bilharz Research Institute under reference (Teodor Bilharz Research Institute REC Protocol No Pt 541).

Preoperative imaging was retrograde urethrogram and antegrade cystourethrogram. Post-operative evaluation of USS-PROM and sexual function was done at 12 months.

Clinician reported outcome was done at 6 and 12 months with uroflowmetry and antegrade urethrogram.

The validated patient-reported outcome measures USS-PROM comprise a LUTS domain and a health-related quality of life domain. The LUTS domain is composed of six items, the questions generate a total score that varies from 0 (asymptomatic) to 24 (most symptomatic). The voiding score ranges between 1 (best) and 4 (worst). The test expresses significant improvements in condition specifics as well as HRQOL components and quantifies changes in voiding symptoms following USS (11),(15). The questionnaire was filled by the patients after 12 months following endo-urethrotomy and Amplatz dilation. Interpretation of USS-PROM was graded into asymptomatic, mildly symptomatic, most symptomatic.

Sexual function (SF) assessment is a questionnaire on pre- and post-treatment erection and ejaculation. We applied the inventory proposed by O’Leary et al (15) for assessment of SF before and after endo-urethrotomy which compares outcomes with normative data. Sexual function was evaluated as: normal SF, weak SF, and sexual dysfunction.

Clinician-derived outcome measurements are maximum flow rate (Qmax) and retrograde urethrogram.

### Surgical procedure

The patient, placed on the operation table in lithotomy position, receives regional anesthesia. The x-ray image intensifier is placed to view the pelvis and perineum. Retrograde urethrogram and antegrade cystogram are done simultaneously to localize the site and length of the stricture area (Figure 1).

**Figure 1.**
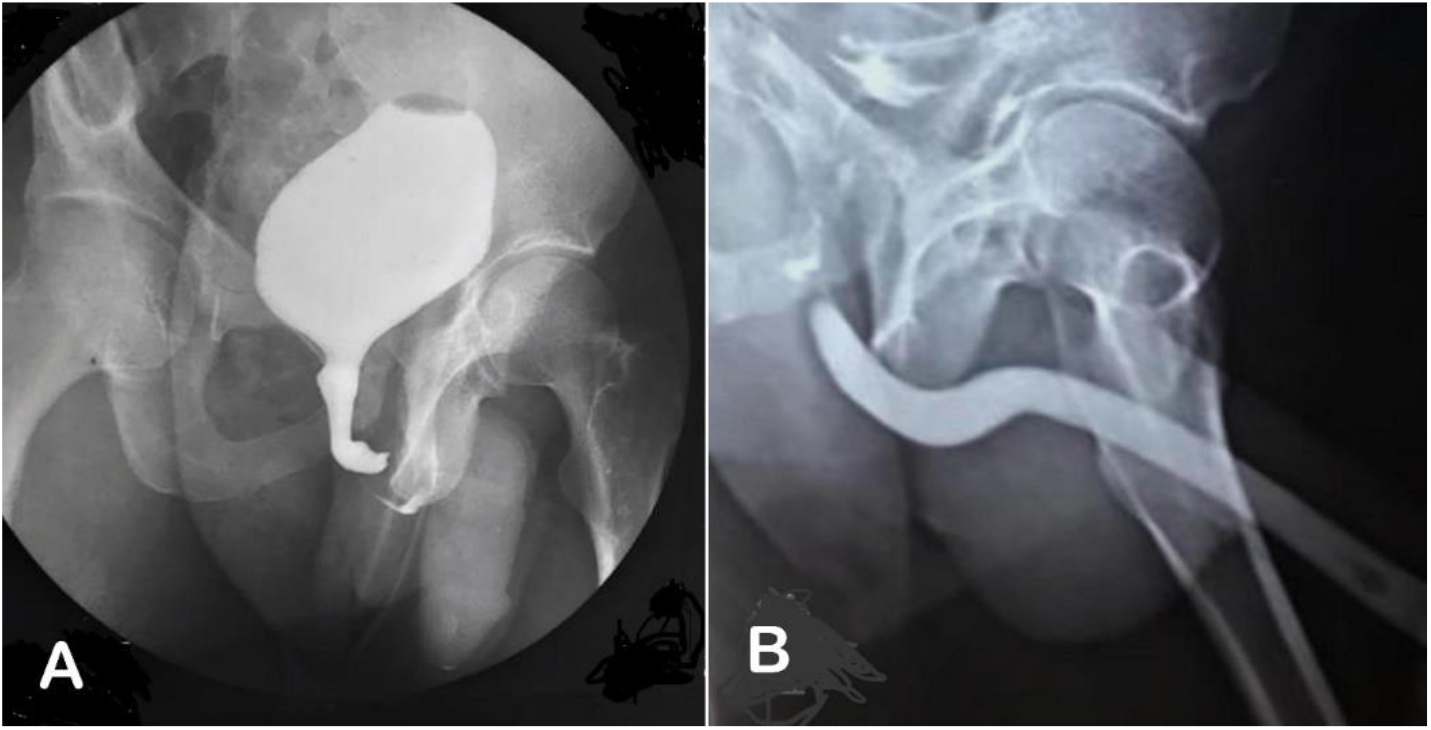
Intra-operative imaging of a patient having posterior urethral stricture showing the site of urethral stricture A) Antegrade cystogram B) Retrograde urethrogram.

A 20 Ch Sachse urethrotome sheath including the diagnostic inner sheath is forwarded in the penile urethra to the site of the stricture. An open end 6 Ch ureteric catheter is passed via the ureteric channel of the inner sheath to the stricture, a hydrophilic guidewire is passed in the ureteric catheter to reach the stricture site and is negotiated under guidance of the imaging intensifier to cross the stricture to reach the bladder, followed with the ureteric catheter.

The hydrophilic guidewire is exchanged with stiff guidewire, the internal sheath is removed and the first Alken metal dilator is passed over the guidewire and negotiated under X-image intensifier to reach the bladder.

Amplatz renal dilators are passed over the Alken dilator one after the other beginning from 6 Ch to 14 Ch. The dilators are removed leaving behind the guidewire.

The Sachse urethrotome with its working element is passed into the penile urethra to the stenotic segment, a cold straight knife is used to make 3 cuts in the stricture segment at 12, 3, 9 points under direct vision. Then, the urethrotome is advanced to enter the bladder, the working element is removed leaving the guidewire in place. The first Alken metal dilator is passed over the guidewire under fluorescent guidance, dilatation is done using Amplatz renal dilators reaching 26-28 Ch. Lastly, a pure silicon indwelling urethral catheter is introduced into the bladder (Figure 2). The suprapubic catheter is to be removed after 3 days and the indwelling catheter after 2 weeks.

**Figure 2.**
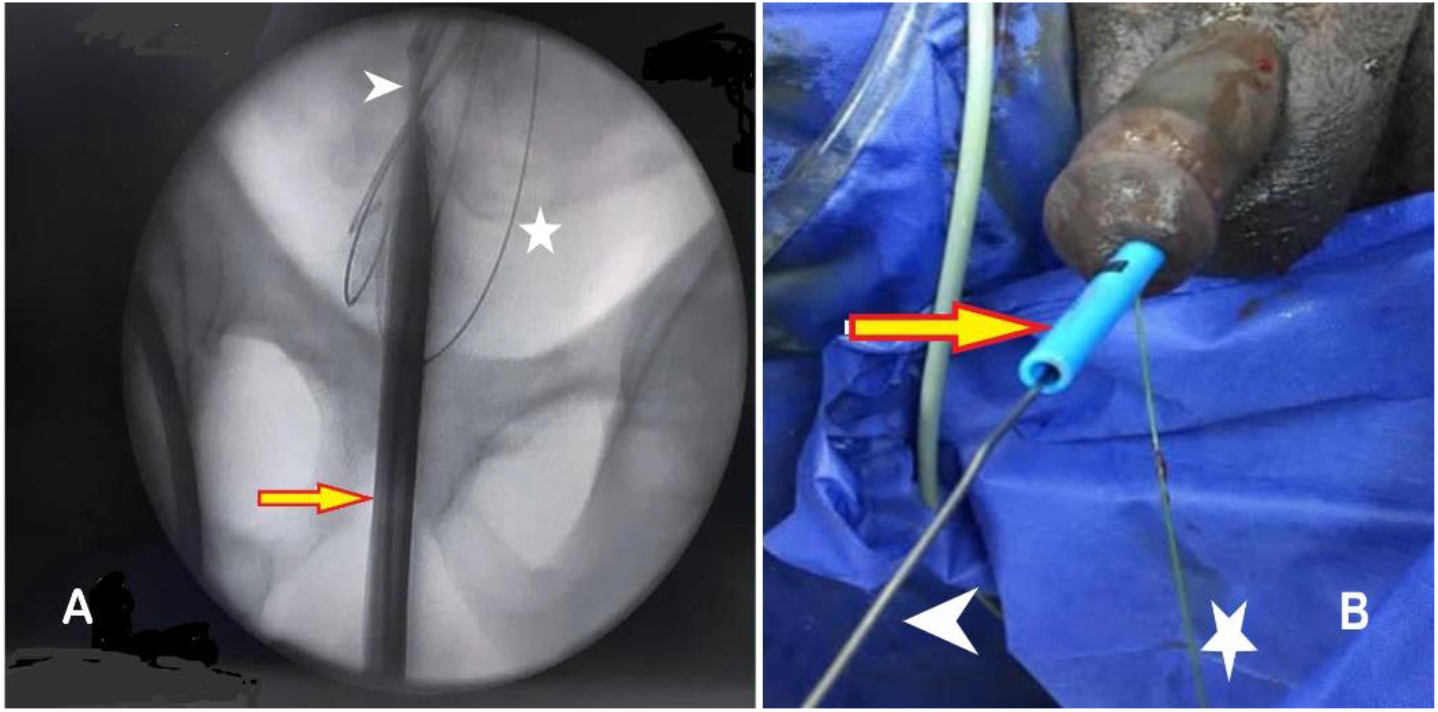
Intra-operative procedure of endourethrotomy. A) X-ray flourescent imaging showing the guidewire (asterix), passed over the first Alken metal renal dilator (arrowhead), and finally the Amplatz renal dilator (arrow) dilated the stricture and advanced to the bladder. B) Operative photograph showing the guidewire (asterix) outside the set of dilator to work as safety guidewire, the Alken metal renal dilator (arrowhead) and Amplatz renal dilator (arrow) are the urethra after dilating the stricture and were advanced to the bladder.

### Ethical consideration

The research followed the tenets of the last edition of Declaration of Helsinki guidelines, eligible patients provided written informed consent, the study was reviewed and approved by the ethics committee of Teodor Bilharz Research Institute under reference (Teodor Bilharz Research Institute REC Protocol No Pt 541).

## RESULTS

The surgical procedures went smoothly. Evaluation of the procedure was done at 6- and 12-months follow-up that showed a patent posterior urethra (Figure 3).

**Figure 3.**
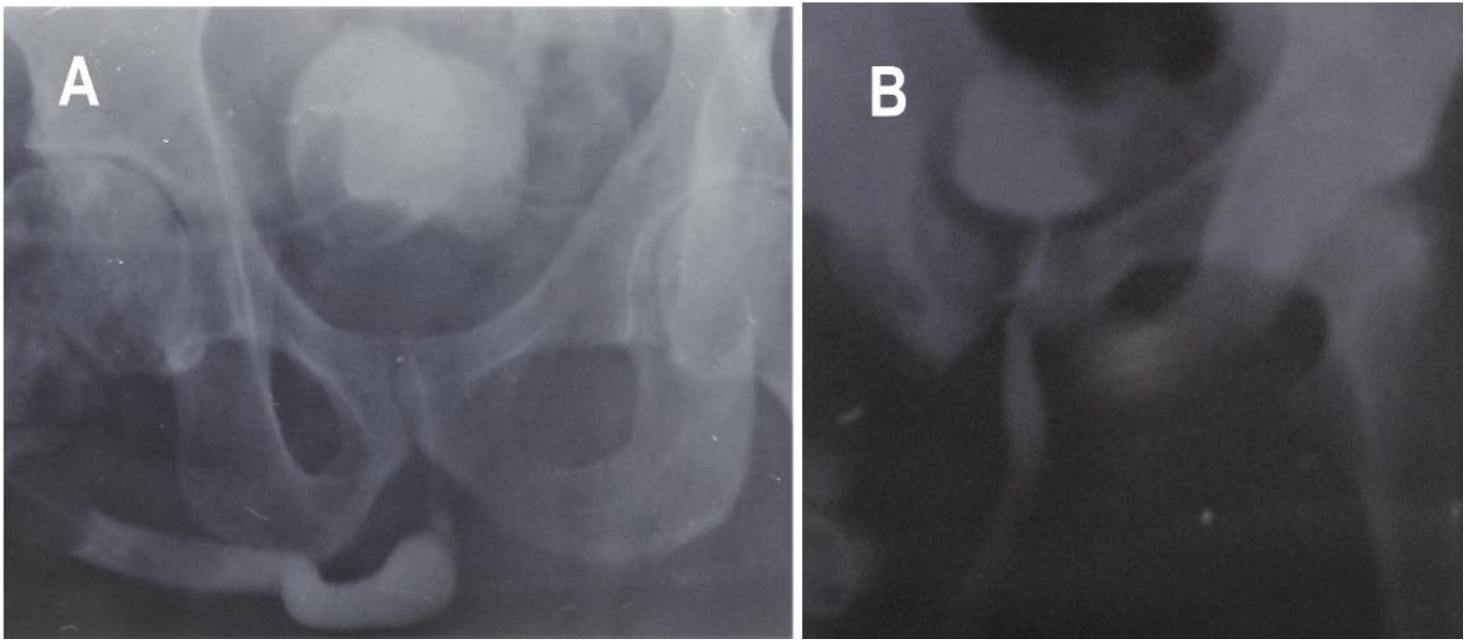
Pre-operative and post-operative imaging in follow-up after endourethrotomy combined with Amplatz dilatation of the stricture area to 28 F. A) Pre-operative simultaneous retrograde urethrogam with antigrade cystograph through suprapubic tube showing the stricture area. B) Post-operative retrograde urethrogram showing patent urethra.

Uroflowmetry at 12 months for the 48 patients showed that mean (minimum - maximum) Qmax was 16.2 (15-20.7) ml/sec.

*Complications* Restenosis after 5 months in one case that was treated by a second endo-urethrotomy; mild stenosis in three patients who were treated with Amplatz dilatation; urinary tract infection in two cases that were treated with urethral dilatation and antibiotics. (Table 2).

**Table2.** complications.

|  | Patients (%) | Symptoms | Qmax (range) | Treatment | Follow-up (12 mos.) |
| --- | --- | --- | --- | --- | --- |
| Re-stricture | 1(2%) | LUTS++++ | 7-9 ml/sec | Endourethrot-omy | Asymptomatic |
| Mild stenosis | 3(6%) | LUTS++ | 10-13 ml/sec | Amplatz dilatation | Asymptomatic |
| Recurrent UTI | 2(4%) | LUTS ++ | 11-15 ml/sec | Bougee dilatation | Asymptomatic |

LUTS: Lower urinary tract symptoms; UTI: urinary tract infection
Total complications 12.5%.

The results of the 48 patients were categorized into 3 groups: I, II, III, according to age as mean. year. (range). The groups represented young, middle, and older ages. This classification aimed at considering the difference in status of LUTS as well as sexual function before the trauma and the prevalence of comorbidities.

Patient-reported outcome was expressed by answering a questionnaire on USS-PROM and sexual function. At 12 months follow-up, the young and middle age groups reported asymptomatic for USS-PROSM and normal for sexual function. The older age group reported asymptomatic in 80%, mildly symptomatic 10%, and most symptomatic in 10 percent. The patients who reported mildly symptomatic and most symptomatic had multiple comorbidities (Table 3).

**Table3.** Postoperative parameters at 12 months follow-up.

| **Table3 Postoperative parameters at 12 months follow-up** |  |  |  |
| --- | --- | --- | --- |
|  | **Group I**<br>**(n=21)** | **Group II**<br>**(n=17)** | **Group III**<br>**(n=10)** |
| Age. median. Years (range) | 23(18-50) | 54(50-65) | 67(65-77) |
| Co-morbidity(CVD,DM,COPD) |  |  |  |
| Single | None | 1 | 2 |
| Multiple | None | 1 | 3 |
| USS-PROM |  |  |  |
| Asymptomatic. no. (%) | 21<br>(100%) | 15<br>(88.2%) | 8 (80%) |
| Mildly symptomatic. no. (%) | 0 (0%) | 2 (11.7%) | 1 (10%) |
| Most symptomatic. no. (%) | 0 (0%) | 0 (0%) | 1 (10%) |
| Sexual function(SF) |  |  |  |
| Normal SF. no. (%) | 21(100%) | 11(100%) | 6 (60%) |
| Weak SF. No. (%) | ===== | ===== | 1 (20%) |
| Sexual dysfunction. no. (%) | ===== | ===== | 3 (30%) |
| Uroflowmetry (Qmax) |  |  |  |
| Mean | 17.5<br>ml/sec | 16.7<br>ml/sec | 16.2 ml/sec |
| Minimum-maximum | 18.6-20.7<br>ml/sec | 16.8-17.5<br>ml/sec | 15-17.3 ml/sec |
| Retrograde urethrogram | Patent<br>urethra | Patent<br>urethra | Patent urethra |
| CVD= cardiovascular diseases; DM=diabetes mellitus. |  |  |  |
| COPD= chronic obstructive lung disease |  |  |  |

## DISCUSSION

The study showed that the procedure of endo-urethrotomy was successful as shown by evaluation of retrograde urethrogram, uroflowmetry, and patients reported outcome regrading USS-PROMs and sexual function.

In 2020, Goulao et al published their work on surgical treatment for recurrent bulbar urethral stricture in a randomized open-label superiority trial of open urethroplasty versus endoscopic urethrotomy to compare the effectiveness of both procedures. In the primary analysis after 24 months, it was concluded that both urethroplasty and urethrotomy improved voiding symptoms and resulted in similar and better symptom scores (3).

In a randomized study, Osman et al demonstrated no significant difference in recurrence between DVIU and urethral dilatation. The recurrence rate at 12 months was correlated with the length of the stricture (4).

Farrell el al reported on a series of 37 patients having recurrent bulbar urethral stricture or bladder neck contracture and underwent DVIU followed by clean intermittent catheterization. Mean stricture length was 2.0 cm. It was concluded that DVIU provides a minimally invasive and widely available tool to manage complex recurrent urethral strictures <3 cm without significant morbidity (5).

In the present series, cold knife urethrotomy was used with good results. Holmium laser was not used. Our results match with those of Jhanwar et al who found that both holmium laser and cold knife urethrotomy are safe and equally effective in treating short-segment bulbar urethral strictures in terms of outcome, complication rate, patient-reported outcome, clinical evaluation in long term follow-up (6).

Endo-urethrotomy begins with the guided dilatation of the stricture under image intensifier followed by DVIU and finally by dilatation with Amplatz renal dilator up to 26 Ch. This procedure ensures proper tract cutting and dilatation. Our procedure is different from that of Nomikos et al who reported success of DVIU following dilatation of complex urethral stricture with Amplatz renal dilators in 34 patients (7).

Akkoc et al treated men with urethral stricture by urethral dilatation using Amplatz renal dilators, without urethrotomy, and reported good results (8).

Azab, in a prospective study comparing the treatment of urethral stricture with either Amplatz renal dilators or with cold knife urethrotomy, evaluated patients for Qmax preoperatively and at 12th months postoperatively. The study concluded that Amplatz dilation is a good option as the initial treatment for urethral stricture. Its results support our findings (9).

Our results are in accordance with other studies which report that the guided urethral dilation and internal urethrotomy are safe, short time procedures and offer satisfactory results with the advance in Qmax at 12 months and the absence of recurrence (10).

This study presents patient-reported outcome measures after endo-urethrotomy using USS-PROM and sexual function questionnaires, each patient has his own control. We showed improvement in HRQOL, USS-PROM, and sexual function scores.

All 48 patients reported being very satisfactory or satisfactory with the procedure. Jackson et al demonstrated that patient-reported outcome measure for urethral stricture surgery is the ideal measurement for assessment of the result of urethral surgery because it represents the patient’s personal perception (11).

In this study, patient-reported quality of sexual function in relation to pre-trauma condition showed that all patients reported very satisfactory following the procedure. Our results are in accordance with Chapman et al who demonstrated in a multi-institutional comparative analysis that non-transecting urethroplasty techniques for bulbar strictures reduce sexual dysfunction in comparison to transecting anastomotic bulbar urethroplasty (12).

The good results achieved in the present study regarding sexual function could be attributed to the non-invasive procedure of endo-urethrotomy which is done without dissection of the posterior urethra that may lead to injury of the neurovascular supply of the penis.

In our study, we stratified the 48 patients into three age groups: young age, middle age, and older age. The groups differ in their pre-trauma conditions regarding voiding function, sexual function, and possibly associated comorbidity. The young age group reported better results and reported very satisfactorily for continence and sexual function.

In this study we used uroflowmetry to evaluate endo-urethrotomy outcomes. Erickson et al used uroflowmetry to measure success and to diagnose recurrent stricture after urethral reconstructive operations (14).

The mean Qmax after endo-urethrotomy was ranging from 18.6-20.7 ml/sec in the young and middle age groups, range of Qmax in older age group was 15-17.3 ml/sec. These variation in Qmax could be attributed to different age groups. Patients in group III had a mean age of 67 years. This age group has the probability of having a component of benign prostatic hyperplasia, a long-standing urethral obstruction with detrusor dysfunction, cardiovascular diseases, or a long history of diabetes mellitus.

The present study has a limitation is that the study was not a prospective randomized study comparing endo-urethrotomy with urethroplasty.

## CONCLUSION

Endo-urethrotomny combined with Amplatz dilation for the treatment of short segment posterior urethral stricture showed promising outcome with no morbidity. Patients’ reporting on USS-PROSM and sexual function showed very satisfactory. Uroflowmetry and retrograde urethrogram after 12 months follow-up showed highly satisfactory results. Further studies with bigger sample sizes and prospective studies comparing endo-urethrotomy with urethroplasties are recommended.

## Data Availability

Data are available on request from the corresponding author.

